# GlycoKnow Ovarian: a Glycoprotein-based, Serum Diagnostic to Distinguish Ovarian Cancers from Benign Pelvic Masses

**DOI:** 10.1101/2025.03.10.25323715

**Authors:** Daniel Serie, Kaitlynn Moser, Maurice Wong, Khushbu Desai, Chad Pickering, Gege Xu, Elizabeth Quach, Meghan Grech, Robert C. Bast, Marcia Ciccone, Tri Dinh, Sudarshan K. Sharma, David Crotzer

## Abstract

**Objective:** Blood-based biomarkers offer an unprecedented opportunity to realize the promise of precision medicine in improving diagnostic workflows. Previous peer-reviewed studies have established the association of the circulating glycoproteome with ovarian cancer. Here a glycoproteomic classifier was built, tested, and applied to both internal and external validation cohorts to distinguish malignant from benign pelvic masses.

**Study Design:** Serum samples from healthy patients and patients with pelvic masses were collected from both retrospective biobanks and prospective trials. In total, 38 peptides and glycopeptides were quantified by InterVenn’s targeted mass spectrometry platform. A classifier to predict malignancy was built, locked, and evaluated in a hold-out test set. The locked diagnostic was then evaluated in an internal validation cohort as well as an external validation cohort from UT MD Anderson Cancer Center.

**Results:** LASSO-regularized logistic regression in the training cohort resulted in a locked classifier with 16 features that was evaluated on a hold-out test set, with strong performance in ovarian cancer and benign pelvic masses (AUC=0.909; sensitivity=86.7%; specificity=89.7%). Comparable performance was observed in further validation cohorts: one internal (stage I-II sensitivity=63.6%; stage III-IV sensitivity=85.7%; specificity=82.7%) and one external (stage I-II sensitivity=64.0%; specificity=86.2%), with varying per-stage prevalence.

**Conclusions:** A novel, CA-125-independent glycoprotein panel was developed to help distinguish benign conditions from ovarian cancer. These circulating biomarkers have great potential to detect ovarian cancer while retaining high specificity and could open new avenues for an improved ovarian cancer diagnostic.

**Research Highlights:**

- A glycoprotein-based liquid biopsy demonstrates strong performance in distinguishing ovarian cancer from benign masses
- The locked and validated classifier exhibits comparable performance in multiple validation cohorts
- As an underexplored layer of biology, protein glycosylation has the potential to enable novel precision medicine solutions

## Introduction

In 2025, ovarian cancer was the sixth leading cause of cancer-related deaths in women in the United States [1]. If caught in its earliest stages, ovarian cancer can be effectively treated with surgery and adjuvant chemotherapy [2,3]. Determination of the malignant potential of a pelvic mass can be a challenging task and requires one to interpret symptoms, imaging characteristics, and laboratory data such as tumor markers. Unfortunately, common symptoms of ovarian cancer, such as pelvic pain and abdominal bloating, can be vague and nonspecific, and often do not occur until the cancer is in later stages. As a result, less than 20% of ovarian cancer cases are diagnosed in stage I (when the 5-year survival is 90% or greater), and the majority are diagnosed at later stages where the 5-year survival is at most 40% [4].

Physicians utilize guidelines from the National Comprehensive Cancer Network and the American College of Obstetricians and Gynecologists in their assessment of malignancy. These groups recommend various diagnostic modalities, including imaging and blood-based tests, to assess the proper course of action. Imaging techniques, primarily transvaginal ultrasound and computerized tomography, are used to determine the presence of a mass [5].

The tumor marker CA-125, a protein found on the cell surface and expressed in blood [6], has historically been used to help differentiate benign from malignant masses, but it suffers from low sensitivity in early disease and a lack of specificity [7]. In fact, CA-125 is not increased in 20% of ovarian carcinomas [5], and elevated CA-125 expression has been seen in other cancers and in various non-cancerous indications such as endometriosis and uterine fibroids. As a result, benign masses may be referred to a gynecologic oncologist for specialized care unnecessarily, and malignant masses may inadvertently undergo initial intervention by a surgeon lacking specialized gynecologic oncologic training. In the latter case, this often leads to additional procedures and potentially adverse clinical outcomes for patients.

Several blood-based tests have been developed to improve standard of care, largely tests that measure CA-125 longitudinally or in conjunction with other biomarkers. Notable examples include ROMA and OVA1 [8–10]. While many tests showcase differential capabilities compared to CA-125 alone, none have found broad acceptance, due to various issues [11–13]. In particular, the ROMA test measures CA125 and HE4 and the OVA1 test (OVERA) measures expression levels of five circulating proteins (CA-125, apolipoprotein A1, beta-2 microglobulin, transferrin and prealbumin). Both demonstrate improvement in distinguishing ovarian cancer from benign disease in women previously diagnosed with a pelvic mass, but have not seen significant acceptance by the medical community [14,15]. Consequently, there is an unmet clinical need for improved tests for the early detection of ovarian cancer, and for distinguishing benign from malignant pelvic masses.

Evidence has been accumulating on the role glycans play in the development of cancer, particularly ovarian cancer [16–19]. Expanding upon previous research [20], in this multi-cohort study the serum glycoproteome was mined for an ovarian cancer diagnostic signature, powered by targeted mass spectrometry. A total of 38 peptides and glycopeptides were quantified, and a 16 feature classifier that distinguishes malignancy was identified and validated. This diagnostic also applies well to healthy populations. Thus, multi-analyte diagnostic tests derived from blood-based, glycopeptide biomarkers may be an accurate and effective aid in triaging pelvic masses to appropriate surgeons.

## Materials and Methods

### Human serum samples

Serum samples from ovarian cancer patients (n = 148, **Table 1**), those with a benign neoplasm of the ovary (n = 79), and healthy women (n=70) were acquired from Indivumed (Hamburg, Germany). Samples were collected prior to therapeutic intervention and in accordance with applicable regulations for human subjects’ protection. Informed consent was obtained from all participants. The study was approved by an institutional review board (WCG IRB 20223899). Assessment of benign and malignant tumors was based on histopathological analysis of tissue specimens. Samples were stored at –80°C until tested.

**Table 1.**
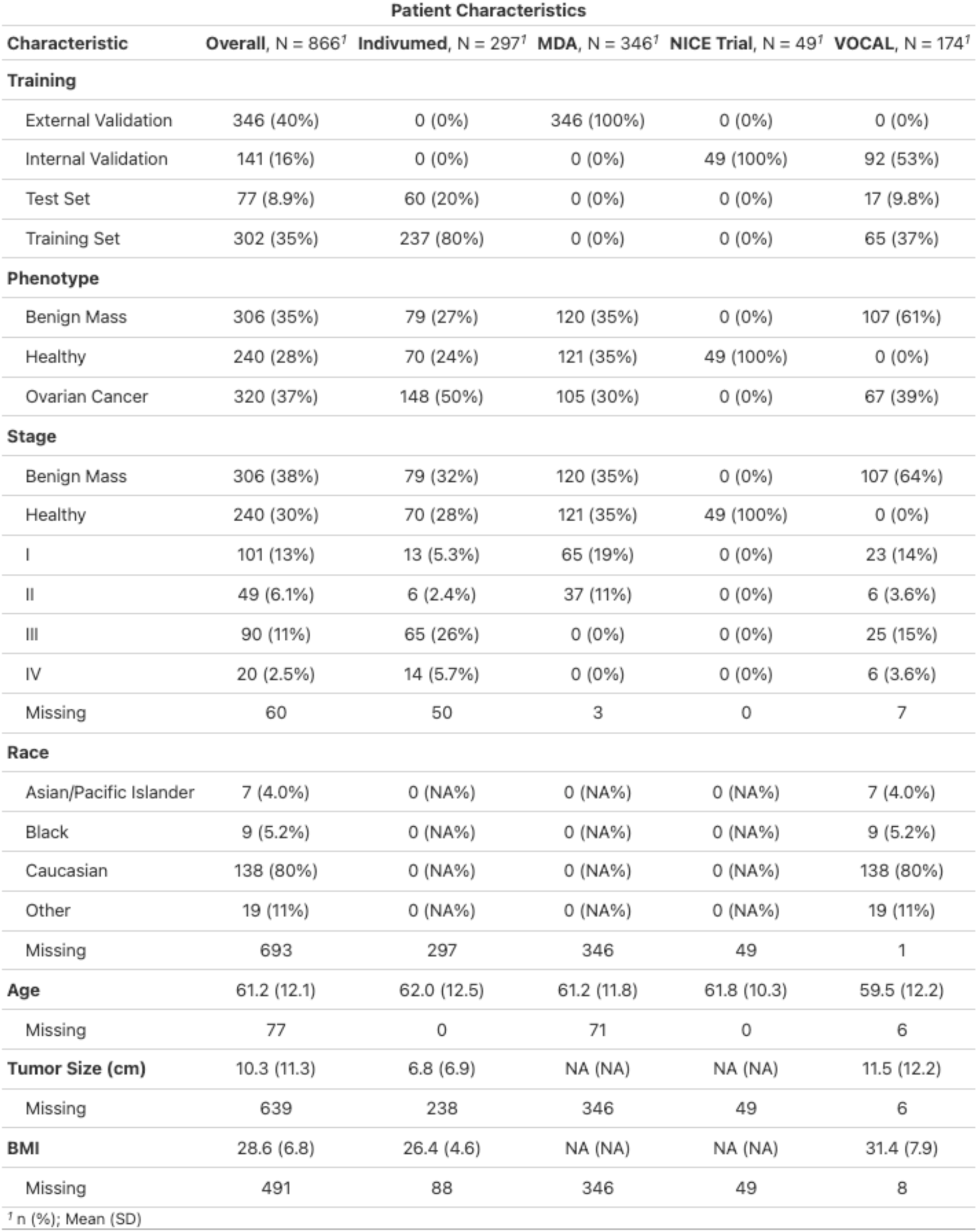
Clinical characteristics of the samples utilized in training and internal validation of the GlycoKnow Ovarian test.

Additionally, this research utilized samples from the InterVenn-sponsored VOCAL trial (NCT03837327), a prospective, international observational study. This IRB-approved study (WIRB IRB 20190246) enrolled women with an imaging-confirmed pelvic mass and prospectively collected biological samples and corresponding clinical, risk factor and histopathology data prior to any biopsy, surgery or intervention. Participants consented to baseline and follow-up data and biospecimen collections, and serum was stored at –80°C until tested. A subset of 82 samples (n = 23 OvCa, n = 59 Benign, **Table 1**) collected in the United States were utilized in the training and testing of the algorithm, and an additional 92 (n = 44 OvCa, n = 48 Benign, **Table 1**) were later selected and employed as an internal validation cohort, to assess performance metrics of the locked classifier.

Furthermore, internal validation was performed on a cohort (n=49, **Table 1**) that had been negatively-screened for adenomas and CRC in the InterVenn-sponsored NICE trial (NCT05445570), an IRB-approved (WIRB IRB 20221935), prospective study in patients seen at colonoscopy clinics. All participants provided written informed consent. Eligible participants aged 45-85 years were identified from patients scheduled for a colonoscopy as part of their standard of care. Individuals with any known malignancy were excluded from the study. Subjects consented to provide a baseline biospecimen collection prior to their scheduled colonoscopy. Clinical data were collected including demographics, medical history, and the pathology report.

External validation samples were acquired in collaboration with MD Anderson under DOD grant <u>(OC190129, Grant ID GRANT12910198)</u>. Serum samples were provided to InterVenn from the MD Anderson prospective clinical study entitled Normal Risk Ovarian Screening Study (NROSS). These samples were divided into two phases (n = 346 total, **Table 1**) and assessed via the same locked classifier.

### Chemicals and reagents

Human serum, dithiothreitol (DTT), and iodoacetamide (IAA) were purchased from MilliporeSigma (St. Louis, MO, USA). Sequencing-grade trypsin was purchased from Promega (Madison, WI, USA), LC-MS-grade water and acetonitrile were obtained from Honeywell (Muskegon, MI, USA), and LC-MS grade formic acid was acquired from Thermo Scientific (Waltham, MA, USA). Stable isotope-labeled peptide internal standards were purchased from Biosynth (formerly Vivitide, Gardner, MA).

### Sample preparation and LC-MS analysis

Serum samples were first treated with DTT and IAA to reduce and alkylate disulfide bonds, which was followed by digestion with trypsin at 37°C for 18 hours. The digestion was quenched by adding formic acid to a final concentration of 1% (v/v). Then finally a cocktail of stable isotope-labeled peptide internal standards was added at known concentrations. Digested serum samples were separated over a Waters ACQUITY UPLC Peptide HSS T3 C18 column (2.1 mm x 50 mm i.d., 1.8 μm particle size) using an Agilent 1290 Infinity UHPLC system. The mobile phase A consisted of 0.1% formic acid in water (v/v), and the mobile phase B of 0.1% formic acid in acetonitrile (v/v), with the flow rate set at 0.4 mL/minute. Analytes were separated with mobile phase B increasing from 1% to 44% over an 18-min elution gradient. After electrospray ionization in positive ion mode, peptides and glycopeptides in the samples were introduced into an Agilent 6495 triple quadrupole MS operated in dynamic multiple reaction monitoring (dMRM) mode for targeted quantification. Samples were injected in a randomized order with respect to the clinical features and phenotypes and interspersed with pooled reference samples.

### Glycopeptide quantification and normalization

PeakBoundaryNet [21] was used to integrate chromatogram peaks and to obtain molecular abundance quantification for each peptide and glycopeptide. R Libraries ‘stats’ and ‘caret’ were used for all statistical analyses and for building machine learning models. In total, 38 peptides and glycopeptides deriving from 19 serum glycoproteins were quantified. Intensity of glycopeptides and peptides were corrected for between-batch drift using reference samples. Raw abundance of peptides was normalized by using heavy isotope-labeled internal standards with known concentrations to determine peptide concentration. Relative abundance of a glycopeptide was calculated as the ratio of the raw abundance of the glycopeptide to the raw abundance of a non-glycosylated peptide from the same glycoprotein. Site occupancy was calculated as the ratio of the raw abundance of any given glycopeptide to the sum of raw abundances of all glycopeptides with the same amino acid sequence. Approximate glycopeptide concentration was calculated by multiplying relative abundance or site occupancy of a glycopeptide by the concentration of a peptide from the same protein. Multiple normalization schemes were employed, as they represent different biological hypotheses. For instance, biomarkers may show associations at the level of absolute concentration that would not be reflected in the percentage of overall glycoforms, and vice-versa. Log-transformed peptide concentration, glycopeptide concentration, site occupancy, and relative abundance were used in modeling steps, totaling 43 unique features.

### Statistical analyses

Fold changes for individual peptides and glycopeptides were calculated in the training and test cohorts based on normalized abundances, comparing ovarian cancer vs controls (benign pelvic mass or healthy). Healthy controls were employed alongside the benign patients to augment the power of the classifier as well as to ensure appropriate results in the greater population of women without pelvic masses. To compare the two phenotypes, age-adjusted linear regression was used on a feature-by-feature basis with phenotype serving as the sole binary independent variable. Differences of any biomarker among phenotype groups compared were considered statistically significant if a p-value of less than 0.05 was reached in both prospective and retrospective cohorts.

Prior to multivariable modeling, normalized data were scaled such that the distributions of all input features were Gaussian with zero mean and unit variance. Patients from Indivumed and the initial VOCAL cohort were split into 80% training (n = 302) and 20% test (n = 77) sets such that phenotype and age were balanced. Ten-fold repeated cross-validation was employed in the training set to select optimal hyperparameters for a LASSO-regularized logistic regression model built on the complete training data. This model was locked and applied to the holdout test sets. Means and standard deviations from the training data were applied to scale internal and external validation cohorts, and the same locked model was applied to these to assess performance in these independent data sets. Sensitivity and specificity were reported separately across phenotypes and cancer stages in cohort, to compare performance across populations with varying distributions of patients. In the ultimate model an indeterminate range of ∼10% of patients where the classifier did not distinguish conditions well, was established in the training cohort [22].

## Results

### Varied cohort demographics represent a broad swath of the population of women with pelvic masses

In total, 174 women from the VOCAL trial, 49 women from the NICE trial, 297 women from the Indivumed biobank, and 346 samples from MD Anderson were analyzed to build and evaluate the test (n=866, **Table 1 and Supplementary Table 1**).

In the VOCAL trial, mean age was similar in the across training and validation sets, ranging from 59.5 to 62.0 (**Table 1**), respectively. Prevalence of malignancy in pelvic masses enrolled was similar to published estimates of 20%, with the training VOCAL data at 24%, as these samples were cumulatively enrolled prior to a freeze date of April 2021. The majority of trial samples utilized in training or testing cohorts were of epithelial ovarian cancer type (60/66 combined, 90.9), but across cohorts the benign and cancer histologies varied (**Supplementary Table 1)**. Race in the VOCAL cohort was 80% Caucasian.

Indivumed samples had a larger fraction of late-stage cancers (80.6%), to guide machine learning algorithms in establishing the trend amongst more extreme glycoproteomic changes. MD Anderson samples were completely stage I and II, which allowed for an unbiased assessment of the classifier performance in the disease’s earliest stages.

### Serum-based glycoproteomic markers distinguish ovarian malignancies from benign tumors and healthy subjects

Differential expression analysis was performed in the combined training and test cohorts, and fold changes and significance were assessed. Overall, 11 markers differentiated ovarian cancer from benign masses in both the VOCAL trial and in retrospective biobank samples at P < 0.05 and with concordant directions of effect (**Supplemental Table 2**). Of these, one feature represented overall protein concentration, and 10 were site-and-glycan-specific glycopeptides. Notably, five out of the top 10 glycopeptides registered increased sialylation in ovarian cancer patients.

### Training a multivariable classifier

Normalized and scaled peptide and glycopeptide markers from the assay were input into a feature selection and weighting loop. Ten-fold cross-validation was utilized to assess the optimal shrinkage parameter for a LASSO model, which was found to be 0.0408. This led to a model with 16 features derived from 14 glycopeptides (**Table 2**). From this model, AUC values of 0.893 in the training and 0.893 in the test set were observed (**Figure 1a**). These values translate to test set values of 88.2% sensitivity and 81.4% specificity (PPV = 78.9%, NPV = 89.7%). Nine out of the sixteen markers on the resulting panel were fucosylated, consistent with InterVenn’s previously published work in ovarian cancer [20].

**Figure 1.**
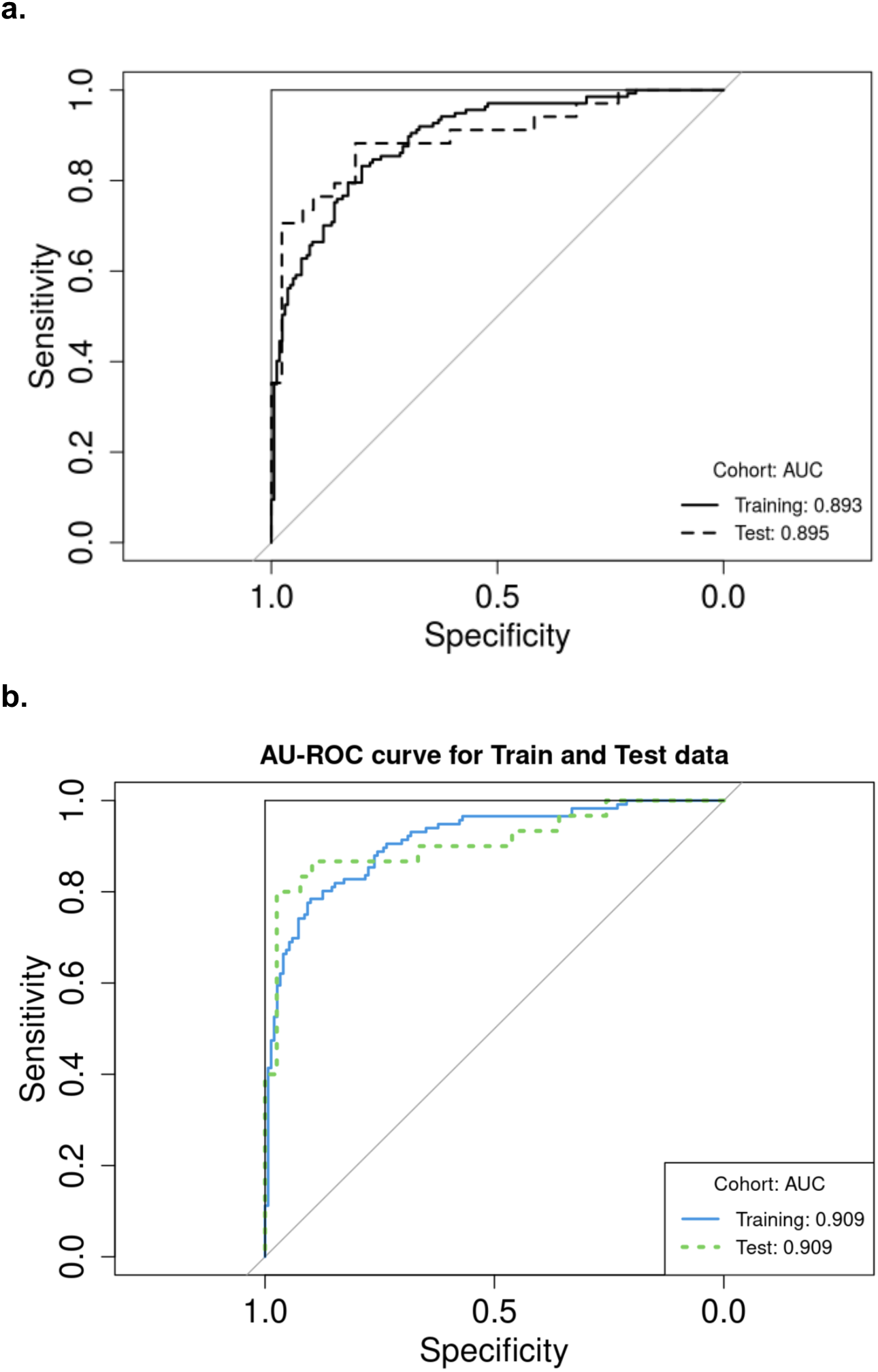
Multivariable analysis reveals a distinct glycoproteomic differentiator of ovarian cancer. **a)** Receiver-Operating Characteristic (ROC) curve representing the sensitivity and specificity of the trained model at different cutoffs, for both training (n=302) and hold-out test (n=77) sets. **b)** ROC curve for the model, removing samples with Indeterminate predictions (removing n=35 in training and n=8 test).

**Table 2.**
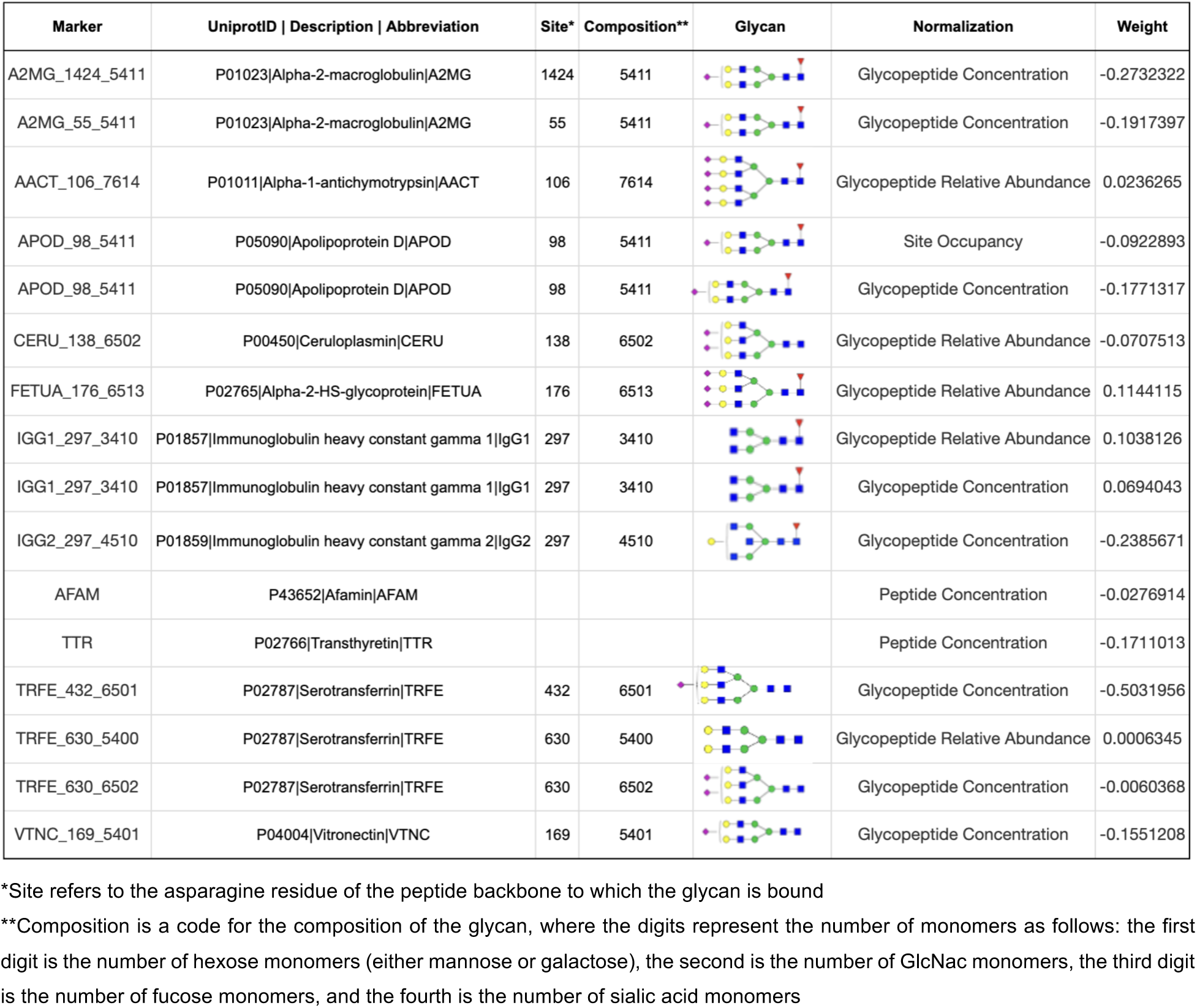
Weights and descriptions of the biomarkers retained in the multivariable model.

| Marker | UniprotID \| Description \| Abbreviation | Site* | Composition** | Glycan | Normalization | Weight |
| --- | --- | --- | --- | --- | --- | --- |
| A2MG_1424_5411 | P01023\|Alpha-2-macroglobulin\|A2MG | 1424 | 5411 |  | Glycopeptide Concentration | -0.2732322 |
| A2MG_55_5411 | P01023\|Alpha-2-macroglobulin\|A2MG | 55 | 5411 |  | Glycopeptide Concentration | -0.1917397 |
| AACT_106_7614 | P01011\|Alpha-1-antichymotrypsin\|AACT | 106 | 7614 |  | Glycopeptide Relative Abundance | 0.0236265 |
| APOD_98_5411 | P05090\|Apolipoprotein D\|APOD | 98 | 5411 |  | Site Occupancy | -0.0922893 |
| APOD_98_5411 | P05090\|Apolipoprotein D\|APOD | 98 | 5411 |  | Glycopeptide Concentration | -0.1771317 |
| CERU_138_6502 | P00450\|Ceruloplasmin\|CERU | 138 | 6502 |  | Glycopeptide Relative Abundance | -0.0707513 |
| FETUA_176_6513 | P02765\|Alpha-2-HS-glycoprotein\|FETUA | 176 | 6513 |  | Glycopeptide Relative Abundance | 0.1144115 |
| IGG1_297_3410 | P01857\|Immunoglobulin heavy constant gamma 1\|IgG1 | 297 | 3410 |  | Glycopeptide Relative Abundance | 0.1038126 |
| IGG1_297_3410 | P01857\|Immunoglobulin heavy constant gamma 1\|IgG1 | 297 | 3410 |  | Glycopeptide Concentration | 0.0694043 |
| IGG2_297_4510 | P01859\|Immunoglobulin heavy constant gamma 2\|IgG2 | 297 | 4510 |  | Glycopeptide Concentration | -0.2385671 |
| AFAM | P43652\|Afamin\|AFAM |  |  |  | Peptide Concentration | -0.0276914 |
| TTR | P02766\|Transthyretin\|TTR |  |  |  | Peptide Concentration | -0.1711013 |
| TRFE_432_6501 | P02787\|Serotransferrin\|TRFE | 432 | 6501 |  | Glycopeptide Concentration | -0.5031956 |
| TRFE_630_5400 | P02787\|Serotransferrin\|TRFE | 630 | 5400 |  | Glycopeptide Relative Abundance | 0.0006345 |
| TRFE_630_6502 | P02787\|Serotransferrin\|TRFE | 630 | 6502 |  | Glycopeptide Concentration | -0.0060368 |
| VTNC_169_5401 | P04004\|Vitronectin\|VTNC | 169 | 5401 |  | Glycopeptide Concentration | -0.1551208 |

\*Site refers to the asparagine residue of the peptide backbone to which the glycan is bound
\*\*Composition is a code for the composition of the glycan, where the digits represent the number of monomers as follows: the first digit is the number of hexose monomers (either mannose or galactose), the second is the number of GlcNac monomers, the third digit is the number of fucose monomers, and the fourth is the number of sialic acid monomers

### Performance excluding “indeterminates”

Consistent with modern diagnostic paradigms [22], an indeterminate range was established for the locked classifier, for probability outputs from 0.4 to 0.5. This results in an increase in performance, with AUC values of 0.909 in both the training and test cohorts (**Figure 1b**), with concomitant effects on the sensitivity (86.7%) and specificity (89.7%).

### Internal validation of the locked classifier

Internal validation samples were run in duplicate to check both performance and test reproducibility. Two samples had duplicates that did not pass QC, so overall, 280 samples were assessed. Of these, 37 were found to have Indeterminate results. In the remaining 81 ovarian cancer patients, 72.8% sensitivity was observed, and in the 162 controls, 82.7% specificity was achieved (76.2% benign, and 89.7% healthy). Later stage ovarian cancer generally displayed increased sensitivity (Stage I – IV sensitivity = 70.4% / 33.3% / 83.3% / 100%). The test concordance correlation coefficient between duplicates was measured at 0.97 (**Figure 2**).

**Figure 2.**
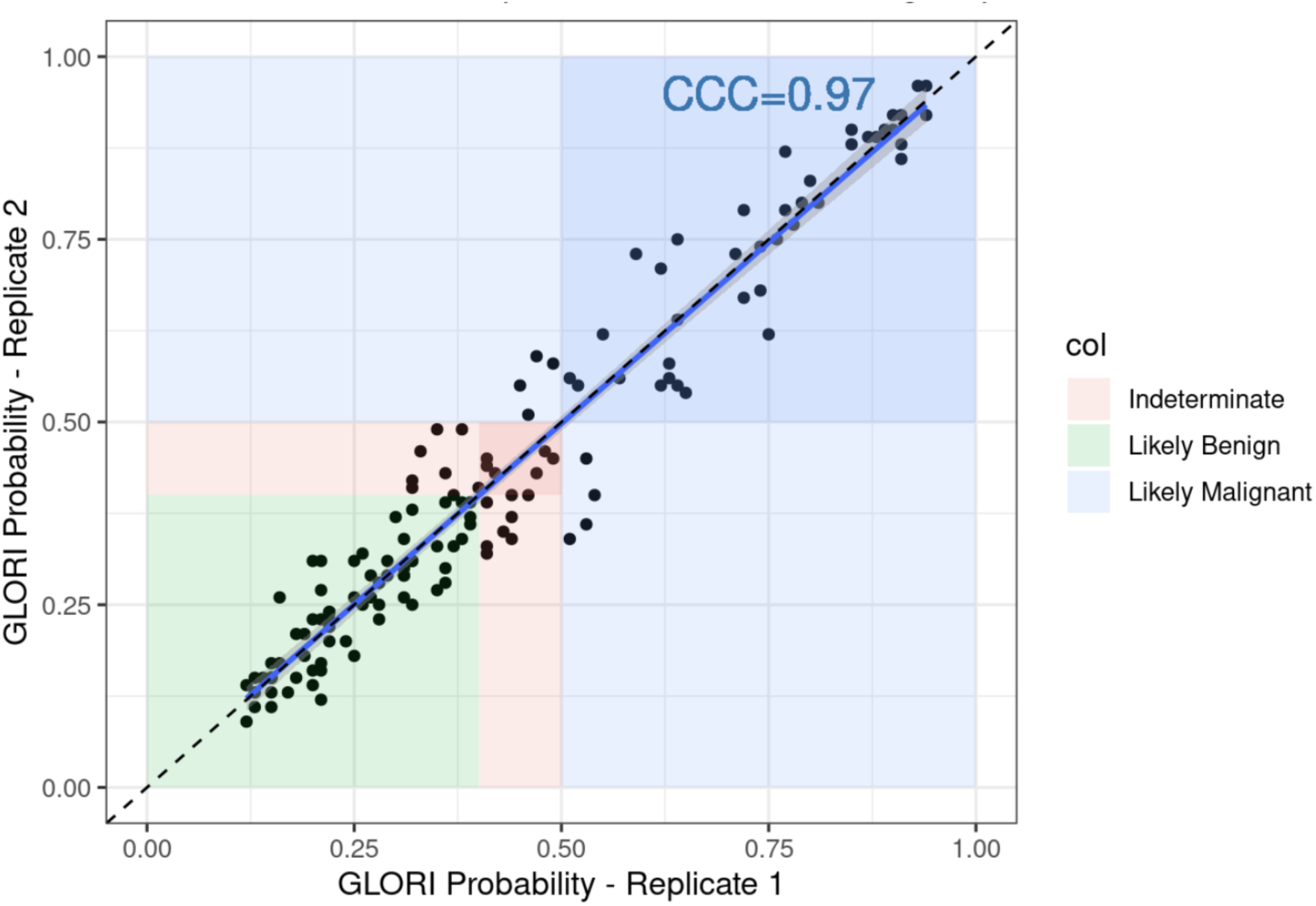
The GlycoKnow Ovarian test displays high reproducibility in replicate analysis. Internal validation samples were run in duplicate to assess reproducibility in the ultimate model output. The concordance correlation coefficient (CCC) was calculated at 0.97.

Notably, while the classifier output correlates with increasing cancer stage, no such association is seen with the size of the pelvic mass. The mass size (in mm) for VOCAL trial samples across both training and internal validation cohorts was compared with probability of malignancy (**Figure 3**). No significant association was observed in benign or malignant masses individually, nor was it observed in the combined set (Spearman correlation 0.174).

**Figure 3.**
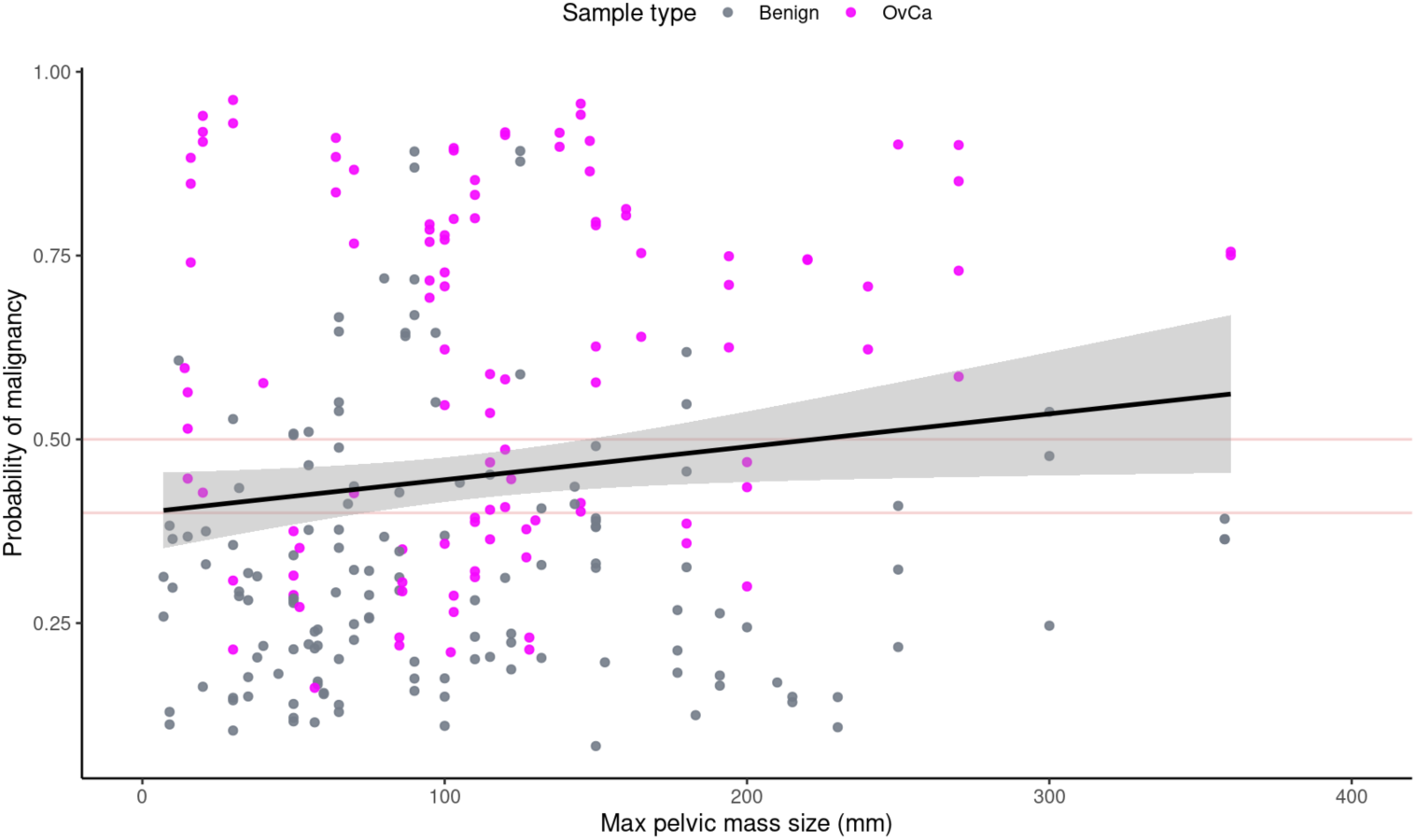
The probability of malignancy displays no association with size of the pelvic mass. Pelvic mass size (mm) was compared with the GlycoKnow output for all VOCAL trial samples, across training and internal validation sets. The Spearman correlation was 0.174, and no significant association was observed.

### Comparison with CA-125 in VOCAL Trial Samples

To compare GlycoKnow Ovarian performance with commercially available Ovarian Cancer diagnostics, CA-125 was assessed on a modest subset of the samples in the VOCAL trial cohorts (**Supplemental Table 3**). The subset of samples utilized in training and test loops (n=59) registered 66.7% sensitivity and 71.1% specificity (in benign pelvic masses). The US internal validation samples (n=72) saw 73.0% sensitivity and 74.3% specificity; combined performance registered at 72.6% sensitivity and 70.7% specificity.

### External validation

External validation samples in stage I and II ovarian cancer, benign pelvic masses, and healthy women were acquired for an unbiased test of classifier performance in these populations. These samples were run in two phases. Combined, 36 samples were labeled Indeterminate and n=310 were predicted as malignant or benign. Overall the test had 64.0% sensitivity for early stage cancer and 86,2% specificity (75% benign, 96% healthy).

Cross-cohort results are summarized in **Table 3**. Though the overall sensitivity and specificity across cohorts differed due to variable cohort composition (i.e. different percentages of each cancer stage, as well as in healthy versus benign), the per-stage performance was similar in these distinct populations. In the training, testing, internal validation, and external validation cohorts, stage I-II sensitivity was measured at 65% – 33% (n=3) – 64% – 64%, respectively. Stage III sensitivity was similar at 85% – 86% – 83% – NA, and stage IV was 100% – 100% – 100% – NA. Benign performance varied, but was still within range at 81% – 88% – 76% – 75%, and healthy performance was consistently high at 100% – 92% – 90% – 96%.

**Table 3.** Cross-cohort performance metrics of the GlycoKnow Ovarian diagnostic.

|  | **Biobank and VOCAL Trial** | **Biobank and VOCAL Trial** | **VOCAL and NICE Trials** | **MD Anderson (DoD grant)** | **Combined** |
| --- | --- | --- | --- | --- | --- |
|  | Training<br>n=302<br>(35 Indeterminate - 12%) | Testing<br>n=77<br>(8 Indeterminate - 10%) | Internal Validation<br>n=280<br>(37 Indeterminate - 13%) | External Validation<br>n=346<br>(36 Indeterminate - 10%) | Combined Non-Training<br>n=703<br>(81 Indeterminate - 12%) |
| **Sensitivity** | 80% | 87% | 73% | 63% | 71% (205) |
| **Specificity** | 87% | 90% | 83% | 86% | 85% (417) |
| **Per-Stage Accuracy (N)** |  |  |  |  |  |
| **Stage I** | 69% (13) | 50% (2) | 70% (27) | 62% (55) | 64% (84) |
| **Stage II** | 57% (7) | 0% (1) | 33% (6) | 68% (34) | 61% (41) |
| **Stage I + II** | 65% (20) | 33% (3) | 64% (33) | 64% (89) | 63% (125) |
| **Stage III** | 85% (46) | 86% (14) | 83% (36) | - | 84% (50) |
| **Stage IV** | 100% (12) | 100% (2) | 100% (6) | - | 100% (8) |
| **Stage Unknown** | 76% (38) | 100% (11) | 50% (8) | 33% (3) | 73% (22) |
| **Benign** | 81% (98) | 88% (26) | 76% (82) | 75% (106) | 77% (214) |
| **Healthy** | 100% (53) | 92% (13) | 90% (78) | 96% (112) | 94% (203) |

### Comment

Ovarian cancer is a complex disease that requires careful triaging and intervention. The most widely used diagnostic laboratory test, CA-125, suffers from suboptimal sensitivity in early-stage disease and inadequate specificity [8]. While newer combinations of biomarkers such as ROMA and OVA-1 perform better than CA-125 alone, novel biomarkers may be necessary to increase utility and uptake in the clinical setting.

### Principal Findings

This work establishes the utility of a targeted panel of circulating glycoproteins as a diagnostic tool to predict the risk of malignancy in pelvic masses. This tool is unique in that it does not incorporate CA-125 but relies on a panel of glycopeptide biomarkers which can predict malignancy, making this an attractive adjunct to CA-125.

This study utilized a robust training cohort, achieving sensitivity and specificity characteristics that outperformed reported sensitivity and specificity values for CA-125 in the literature [8]. When the locked 16 marker classifier was tested against the hold-out set from the VOCAL trial, sensitivity results were high for malignant masses and were particularly robust in the setting of advanced disease. In all cohorts, GlycoKnow specificity results were high. Lastly, GlycoKnow continued to perform well in both internal and external validation cohorts, with varied sources and stage distributions. In the external validation cohort, sensitivity of 64% was seen in early-stage disease, roughly on par with CA-125’s early-stage sensitivity.

While these results demonstrate a strong correlation between disease stage and the performance characteristics of GlycoKnow as a diagnostic tool, the size of pelvic masses does not influence results of the GlycoKnow test (**Figure 3**). This is beneficial from a clinical standpoint, since the size of a mass often influences the decision to refer a patient to a gynecologic oncologist, even though size is often not a reliable indicator of malignancy or stage.

### Clinical Implications

From a clinical perspective, identification and treatment of early-stage cancers has the greatest potential to impact long term survival rates. Current liquid biopsy paradigms largely focus on circulating tumor or cell-free DNA and methylation markers which arise from the tumor itself [23–25]. Accordingly, such targets may be difficult to detect in early-stage disease where biomarker levels might be low. Similarly, CA-125 is a glycoprotein shed from ovarian cancer cells, but not exclusively. Levels can be normal in early stage disease or elevated by a host of benign conditions. In contrast, the glycopeptide biomarkers utilized in this study originate from proteoforms that are secreted by hepatocytes and B-cells. As such, they are more indicative of the body’s response to the presence of cancer rather than the extent of cancer [26]. Utilizing this response mechanism, the GlycoKnow test is able to identify glycoprotein profiles in the presence of early-stage disease and reliably differentiate malignant ovarian cancer from benign conditions.

### Results in the Context of What is Known

The results presented herein expand upon previous studies of ovarian cancer [16–20, 27–29], as a locked glycoproteomic diagnostic has now been applied directly to clinical trial samples. As seen previously [20], fucosylation of proteins was a predominant feature of this ovarian cancer diagnostic, as nine of the sixteen features represented fucosylated glycopeptides. As seen in other investigations [30], sialic acid also plays a prominent role, with ten of the sixteen features containing sialic acid.

### Research Implications

For the most part, the immunoglobulins excepted [18], these specific glycoforms represent novel associations with ovarian cancer. But most of the proteins themselves have been associated with cancer or ovarian cancer previously. Both alpha-2-macroglobulin (A2MG) and transthyretin (TTR) have been identified as diagnosis biomarkers for epithelial ovarian cancer [31]. Alpha1-antichymotrypsin (AACT) has been associated with cancer in general [32], as have apolipoproteins such as APOD [33]. Afamin (AFAM), too, has been linked to ovarian cancer in large patient studies [34]. Transferrin (TRFE) may be linked to ovarian cancer etiology through iron deficiency [35]. And vitronectin (VTNC) itself is responsible for promoting cell invasion and metastasis in a range of cancers [36]. Since the proteomic technologies used to measure these markers do not account for glycosylation, it is perhaps not surprising that specific glycoforms of a protein can display even stronger biological relevance. In spite of tantalizing hints, a definitive mechanism across the proteome remains elusive, and is a target for further research.

### Strengths and Limitations

The strengths of this study included robust training and testing cohorts, as well as both an internal and external validation cohort. In the internal validation cohort, CA-125 sensitivity and specificity results were consistent with reports in the literature, and the sensitivity and specificity values in training, testing and validation cohorts were superior to CA-125 alone. The VOCAL trial dataset, used for testing and internal validation, consisted of prospectively collected samples with corresponding clinical information. The novel panel of 16 biomarkers, the first to mine the circulating glycoproteome for a signature and apply it to multiple trial cohorts, was predictive and reproducible.

There are some limitations to the current study. This study does not compare the performance of GlycoKnow directly to currently available testing modalities such as OVA-1 or ROMA. Likewise, the limited number of in-study samples with CA-125 measurements precludes training a joint classifier with glycoproteomic biomarkers. While including a large selection of ovarian histologies is important in establishing a broadly-applicable cancer diagnostic, evaluating differential performance between these specific subtypes will require further study and clinical validation. Lastly, the ovarian cancers obtained via the MD Anderson external validation sample bank consisted entirely of early stage cancers, which resulted in differential performance compared to the VOCAL trial data, which included late stage cancer.

### Conclusions

Understanding the limitations of current diagnostic modalities available in the workup of pelvic masses, this study supports the utility of a glycoprotein panel to help distinguish benign conditions from ovarian cancer. Being able to more accurately identify benign masses from ovarian cancer could assist in more timely, accurate triaging of patients and ensure more patients undergo appropriate surgical intervention in the management of ovarian cancer. This could expedite care and lead to improved clinical outcomes for patients. As such, GlycoKnow has tremendous clinical potential, and deserves further investigation and validation as a diagnostic tool for women with a pelvic mass being considered for surgical exploration.

## Data Availability

Data produced in the present study are available subject to the approval and consent of all authors.

## Acknowledgments

We’d like to thank all of our colleagues at InterVenn Biosciences for their hard work and support.

We are deeply grateful to all the study participants whose willingness to contribute to research made the VOCAL study possible. Your participation is invaluable in advancing the development of diagnostics for ovarian cancer, bringing us closer to better detection and outcomes. We also extend our sincere appreciation to the physicians, study coordinators, and research lab staff for their dedication and expertise to the VOCAL study. In particular we would like to thank Aparna Kamat, James J. Burke II, Albert Pisani, James Frederick Lilja, Karen Finkelstein, and Matthew Anderson. Your hard work and commitment were essential in successfully completing this study and driving meaningful progress in women’s health.

## Authors’ contributions

Conceptualization: DS, MW. Data curation: KM, MW, KD, CP, CS. Formal analysis: DS, KD, CP. Methodology: DS, MW, GX. Project administration: KM, CS, EQ, MG, VOCAL. Validation: EQ, MG, RB, MC, VOCAL. Visualization: DS, KD, CP. Roles/Writing – original draft: DS, DC. Writing – review & editing: all authors.

## Ethics approval and consent to participate

All subjects gave written informed consent to participate. The study was approved by institutional review boards (WCG IRB 20223899 and WIRB IRB 20190246 and WIRB IRB 20221935) and performed in accordance with the Declaration of Helsinki.

---

**Supplemental Table 1.**
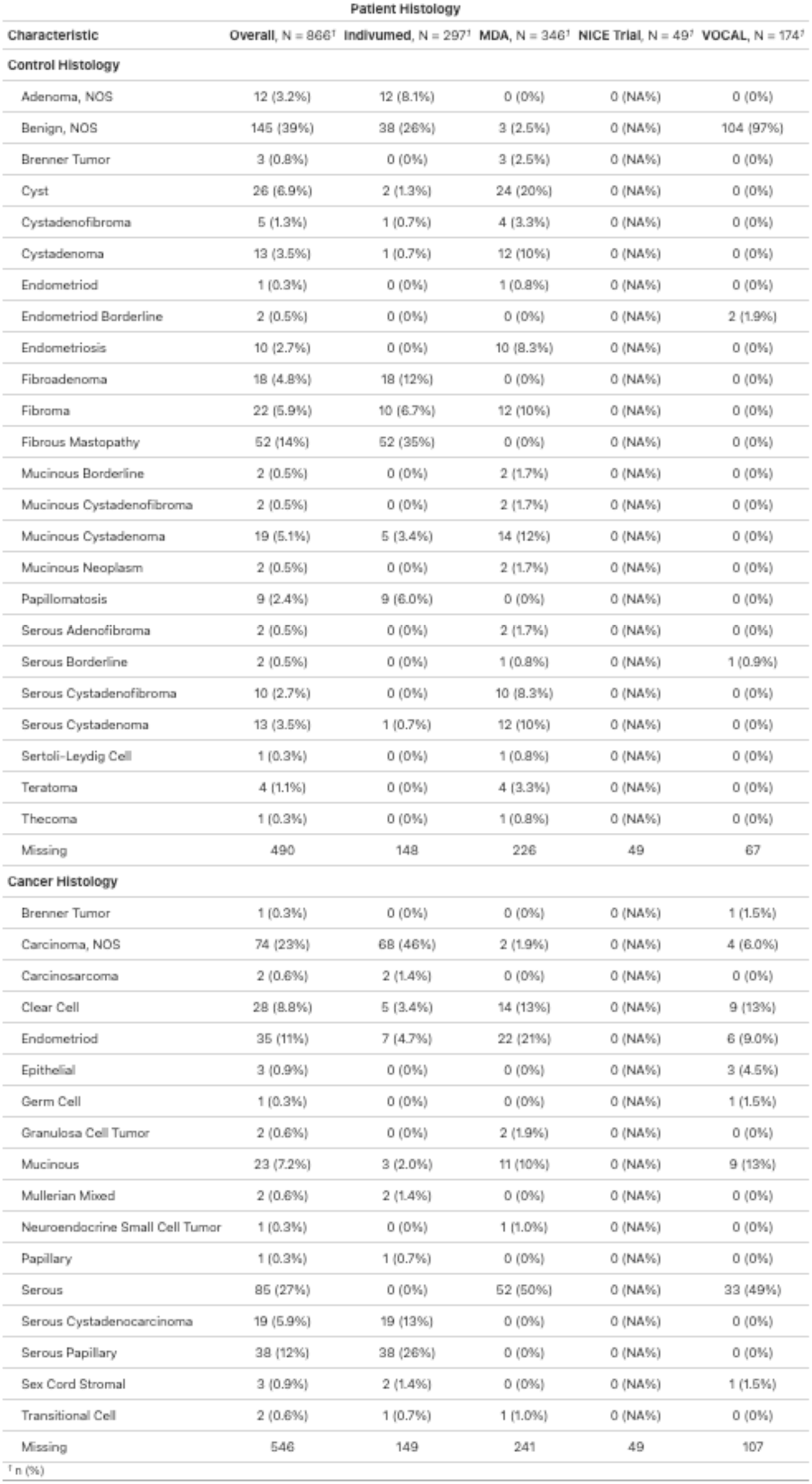
Histological characteristics of all cohorts utilized in the training, testing, and internal/external validation of the GlycoKnow Ovarian test.

| Characteristic | Patient Histology |  |  |  |  |
| --- | --- | --- | --- | --- | --- |
|  | Overall, N = 866<sup>†</sup> | Indivumed, N = 297<sup>†</sup> | MDA, N = 346<sup>†</sup> | NICE Trial, N = 49<sup>†</sup> | VOCAL, N = 174<sup>†</sup> |
| **Control Histology** |  |  |  |  |  |
| Adenoma, NOS | 12 (3.2%) | 12 (8.1%) | 0 (0%) | 0 (NA%) | 0 (0%) |
| Benign, NOS | 145 (39%) | 38 (26%) | 3 (2.5%) | 0 (NA%) | 104 (97%) |
| Brenner Tumor | 3 (0.8%) | 0 (0%) | 3 (2.5%) | 0 (NA%) | 0 (0%) |
| Cyst | 26 (6.9%) | 2 (1.3%) | 24 (20%) | 0 (NA%) | 0 (0%) |
| Cystadenofibroma | 5 (1.3%) | 1 (0.7%) | 4 (3.3%) | 0 (NA%) | 0 (0%) |
| Cystadenoma | 13 (3.5%) | 1 (0.7%) | 12 (10%) | 0 (NA%) | 0 (0%) |
| Endometriod | 1 (0.3%) | 0 (0%) | 1 (0.8%) | 0 (NA%) | 0 (0%) |
| Endometriod Borderline | 2 (0.5%) | 0 (0%) | 0 (0%) | 0 (NA%) | 2 (1.9%) |
| Endometriosis | 10 (2.7%) | 0 (0%) | 10 (8.3%) | 0 (NA%) | 0 (0%) |
| Fibroadenoma | 18 (4.8%) | 18 (12%) | 0 (0%) | 0 (NA%) | 0 (0%) |
| Fibroma | 22 (5.9%) | 10 (6.7%) | 12 (10%) | 0 (NA%) | 0 (0%) |
| Fibrous Mastopathy | 52 (14%) | 52 (35%) | 0 (0%) | 0 (NA%) | 0 (0%) |
| Mucinous Borderline | 2 (0.5%) | 0 (0%) | 2 (1.7%) | 0 (NA%) | 0 (0%) |
| Mucinous Cystadenofibroma | 2 (0.5%) | 0 (0%) | 2 (1.7%) | 0 (NA%) | 0 (0%) |
| Mucinous Cystadenoma | 19 (5.1%) | 5 (3.4%) | 14 (12%) | 0 (NA%) | 0 (0%) |
| Mucinous Neoplasm | 2 (0.5%) | 0 (0%) | 2 (1.7%) | 0 (NA%) | 0 (0%) |
| Papillomatosis | 9 (2.4%) | 9 (6.0%) | 0 (0%) | 0 (NA%) | 0 (0%) |
| Serous Adenofibroma | 2 (0.5%) | 0 (0%) | 2 (1.7%) | 0 (NA%) | 0 (0%) |
| Serous Borderline | 2 (0.5%) | 0 (0%) | 1 (0.8%) | 0 (NA%) | 1 (0.9%) |
| Serous Cystadenofibroma | 10 (2.7%) | 0 (0%) | 10 (8.3%) | 0 (NA%) | 0 (0%) |
| Serous Cystadenoma | 13 (3.5%) | 1 (0.7%) | 12 (10%) | 0 (NA%) | 0 (0%) |
| Sertoli-Leydig Cell | 1 (0.3%) | 0 (0%) | 1 (0.8%) | 0 (NA%) | 0 (0%) |
| Teratoma | 4 (1.1%) | 0 (0%) | 4 (3.3%) | 0 (NA%) | 0 (0%) |
| Thecoma | 1 (0.3%) | 0 (0%) | 1 (0.8%) | 0 (NA%) | 0 (0%) |
| Missing | 490 | 148 | 226 | 49 | 67 |
| **Cancer Histology** |  |  |  |  |  |
| Brenner Tumor | 1 (0.3%) | 0 (0%) | 0 (0%) | 0 (NA%) | 1 (1.5%) |
| Carcinoma, NOS | 74 (23%) | 68 (46%) | 2 (1.9%) | 0 (NA%) | 4 (6.0%) |
| Carcinosarcoma | 2 (0.6%) | 2 (1.4%) | 0 (0%) | 0 (NA%) | 0 (0%) |
| Clear Cell | 28 (8.8%) | 5 (3.4%) | 14 (13%) | 0 (NA%) | 9 (13%) |
| Endometriod | 35 (11%) | 7 (4.7%) | 22 (21%) | 0 (NA%) | 6 (9.0%) |
| Epithelial | 3 (0.9%) | 0 (0%) | 0 (0%) | 0 (NA%) | 3 (4.5%) |
| Germ Cell | 1 (0.3%) | 0 (0%) | 0 (0%) | 0 (NA%) | 1 (1.5%) |
| Granulosa Cell Tumor | 2 (0.6%) | 0 (0%) | 2 (1.9%) | 0 (NA%) | 0 (0%) |
| Mucinous | 23 (7.2%) | 3 (2.0%) | 11 (10%) | 0 (NA%) | 9 (13%) |
| Mullerian Mixed | 2 (0.6%) | 2 (1.4%) | 0 (0%) | 0 (NA%) | 0 (0%) |
| Neuroendocrine Small Cell Tumor | 1 (0.3%) | 0 (0%) | 1 (1.0%) | 0 (NA%) | 0 (0%) |
| Papillary | 1 (0.3%) | 1 (0.7%) | 0 (0%) | 0 (NA%) | 0 (0%) |
| Serous | 85 (27%) | 0 (0%) | 52 (50%) | 0 (NA%) | 33 (49%) |
| Serous Cystadenocarcinoma | 19 (5.9%) | 19 (13%) | 0 (0%) | 0 (NA%) | 0 (0%) |
| Serous Papillary | 38 (12%) | 38 (26%) | 0 (0%) | 0 (NA%) | 0 (0%) |
| Sex Cord Stromal | 3 (0.9%) | 2 (1.4%) | 0 (0%) | 0 (NA%) | 1 (1.5%) |
| Transitional Cell | 2 (0.6%) | 1 (0.7%) | 1 (1.0%) | 0 (NA%) | 0 (0%) |
| Missing | 546 | 149 | 241 | 49 | 107 |

<sup>†</sup> n (%)

**Supplemental Table 2.** Significantly differentially expressed markers in both the US VOCAL and Indivumed sample sets.

| Uniprot ID \| Description \| Abbreviation | Site | Composition | FC.All | P.All | FC.US | P.US | FC.IND | P.IND |
| --- | --- | --- | --- | --- | --- | --- | --- | --- |
| P02787\|Serotransferrin\|TRFE | 432 | 6501 | 0.73 | 6.64E-19 | 0.71 | 1.77E-04 | 0.65 | 7.81E-15 |
| P02766\|Transthyretin\|TTR |  |  | 0.70 | 7.47E-20 | 0.66 | 2.69E-04 | 0.67 | 1.40E-14 |
| P04004\|Vitronectin\|VTNC | 169 | 5401 | 0.67 | 7.71E-28 | 0.70 | 5.61E-04 | 0.62 | 7.08E-20 |
| P02787\|Serotransferrin\|TRFE | 630 | 5400 | 1.66 | 1.53E-19 | 1.62 | 6.06E-03 | 1.90 | 1.61E-16 |
| P02763\|Alpha-1-acid glycoprotein 1\|AGP1 | 93 | 7614 | 2.42 | 6.31E-25 | 1.78 | 9.90E-03 | 2.68 | 3.91E-15 |
| P02765\|Alpha-2-HS-glycoprotein\|FETUA | 176 | 6513 | 1.77 | 4.75E-26 | 1.49 | 1.34E-02 | 2.11 | 1.92E-20 |
| P02787\|Serotransferrin\|TRFE | 630 | 6502 | 0.79 | 1.14E-14 | 0.84 | 1.64E-02 | 0.74 | 2.61E-11 |
| P02790\|Hemopexin\|HEMO | 453 | 5402 | 1.05 | 4.16E-20 | 1.04 | 2.17E-02 | 1.07 | 8.05E-18 |
| P01857\|Immunoglobulin heavy constant gamma 1\|IgG1 | 297 | 5510 | 0.67 | 1.79E-18 | 0.74 | 3.04E-02 | 0.59 | 1.30E-14 |
| P02787\|Serotransferrin\|TRFE | 432 | 6503 | 1.35 | 1.44E-21 | 1.17 | 4.42E-02 | 1.49 | 5.72E-19 |
| P01009\|Alpha-1-antitrypsin\|A1AT | 107 | 6513 | 1.46 | 1.13E-10 | 1.27 | 4.52E-02 | 1.64 | 4.78E-09 |
| P02787\|Serotransferrin\|TRFE | 432 | 6502 | 0.81 | 1.24E-14 | 0.86 | 5.35E-02 | 0.73 | 3.26E-14 |
| P02750\|Leucine-richAlpha-2-glycoprotein\|A2GL |  |  | 1.80 | 1.02E-24 | 1.34 | 5.80E-02 | 2.10 | 1.26E-19 |
| P01011\|Alpha-1-antichymotrypsin\|AACT | 271 | 6513 | 1.42 | 6.46E-13 | 1.26 | 7.03E-02 | 1.58 | 1.63E-11 |
| P02763\|Alpha-1-acid glycoprotein 1\|AGP1 | 103 | 8704 | 0.83 | 1.43E-11 | 0.86 | 7.77E-02 | 0.77 | 6.70E-13 |
| P06681\|ComplementC2\|CO2 | 621 | 5200 | 1.16 | 8.05E-14 | 1.12 | 7.95E-02 | 1.20 | 4.11E-11 |
| P01859\|Immunoglobulin heavy constant gamma 2\|IgG2 | 297 | 5510 | 0.59 | 6.42E-17 | 0.68 | 8.33E-02 | 0.49 | 1.80E-15 |
| P01011\|Alpha-1-antichymotrypsin\|AACT | 106 | 7614 | 1.62 | 2.36E-14 | 1.29 | 9.02E-02 | 1.81 | 2.74E-13 |
| P43652\|Afamin\|AFAM |  |  | 0.76 | 9.26E-18 | 0.86 | 9.07E-02 | 0.72 | 7.00E-15 |
| P01857\|Immunoglobulin heavy constant gamma 1\|IgG1 | 297 | 3410 | 1.37 | 1.35E-19 | 1.19 | 9.51E-02 | 1.54 | 2.53E-19 |

\*Site refers to the asparagine residue of the peptide backbone to which the glycan is bound
\*\*Composition is a code for the composition of the glycan, where the digits represent the number of monomers as follows: the first digit is the number of hexose monomers (either mannose or galactose), the second is the number of GlcNAc monomers, the third digit is the number of fucose monomers, and the fourth is the number of sialic acid monomers

**Supplemental Table 3.** CA-125 sensitivity and specificity (at cutoff of 35 U/mL) in VOCAL trial samples.

| CA-125 Performance in VOCAL Trial |  |  |  |
| --- | --- | --- | --- |
|  | Training | Validation | Overall |
| **True Negatives** | 27 | 26 | 53 |
| **False Positives** | 11 | 9 | 20 |
| **False Negatives** | 7 | 10 | 17 |
| **True Positives** | 14 | 27 | 41 |
| **Specificity** | 0.711 | 0.743 | 0.707 |
| **Sensitivity** | 0.667 | 0.73 | 0.726 |

## References

1. Siegel RL , Kratzer TB, Giaquinto AN , Sung H, Jemal A. Cancer statistics, 2025. CA Cancer J Clin. 2025; 75(1): 10–45. DOI:10.3322/caac.21871.

2. Lheureux S, Gourley C, Vergote I, Oza AM. Epithelial ovarian cancer. The Lancet. 2019 Mar 23;393(10177):1240–53.

3. Matulonis UA, Sood AK, Fallowfield L, Howitt BE, Sehouli J, Karlan BY. Ovarian cancer. Nat Rev Dis Primer. 2016 Aug 25;2(1):1–22.

4. Lu KH. Screening for Ovarian Cancer in Asymptomatic Women. JAMA. 2018 Feb;13;319(6):557.

5. Bullock B, Larkin L, Turker L, Stampler K. Management of the Adnexal Mass: Considerations for the Family Medicine Physician. Front Med. 2022 Jul 5;9:913549.

6. Felder M, Kapur A, Gonzalez-Bosquet J, Horibata S, Heintz J, Albrecht R, et al. MUC16 (CA125): tumor biomarker to cancer therapy, a work in progress. Mol Cancer. 2014 May 29;13(1):129.

7. Charkhchi P, Cybulski C, Gronwald J, Wong FO, Narod SA, Akbari MR. CA125 and Ovarian Cancer: A Comprehensive Review. Cancers. 2020 Dec;11;12(12):3730.

8. Dochez V, Caillon H, Vaucel E, Dimet J, Winer N, Ducarme G. Biomarkers and algorithms for diagnosis of ovarian cancer: CA125, HE4, RMI and ROMA, a review. J Ovarian Res. 2019 Mar 27;12(1):28.

9. Skates SJ. Ovarian Cancer Screening: Development of the Risk of Ovarian Cancer Algorithm (ROCA) and ROCA Screening Trials. Int J Gynecol Cancer. 2012;22(S1):S24–6.

10. Montagnana M, Danese E, Ruzzenente O, Bresciani V, Nuzzo T, Gelati M, et al. The ROMA (Risk of Ovarian Malignancy Algorithm) for estimating the risk of epithelial ovarian cancer in women presenting with pelvic mass: is it really useful? Clin Chem Lab Med. 2011 Mar 1;49(3):521–5.

11. Yurkovetsky ZR, Linkov FY, E Malehorn D, Lokshin AE. Multiple biomarker panels for early detection of ovarian cancer. Future Oncol. 2006 Dec;2(6):733–41.

12. McIntosh M, Anderson G, Drescher C, Hanash S, Urban N, Brown P. Ovarian Cancer Early Detection Claims Are Biased. Clin Cancer Res. 2008 Nov;15;14(22):7574–7574.

13. Ueland FR, Desimone CP, Seamon LG, Miller RA, Goodrich S, Podzielinski I, et al. Effectiveness of a Multivariate Index Assay in the Preoperative Assessment of Ovarian Tumors. Obstet Gynecol. 2011 Jun;117(6):1289.

14. Bristow RE, Smith A, Zhang Z, Chan DW, Crutcher G, Fung ET. Ovarian malignancy risk stratification of the adnexal mass using a multivariate index assay. Gynecol Oncol. 2013;Feb;128(2):252–9.

15. Fritsche HA, Bullock RG. A reflex testing protocol using two multivariate index assays improves the risk assessment for ovarian cancer in patients with an adnexal mass. Int J Gynecol Obstet. 2023; 162: 485–492. doi:10.1002/ijgo.14733

16. Alley WR, Vasseur JA, Goetz JA, Svoboda M, Mann BF, Matei DE, et al. N-linked glycan structures and their expressions change in the blood sera of ovarian cancer patients. J Proteome Res. 2012 Apr 6;11(4):2282–300.

17. Miyamoto S, Stroble CD, Taylor S, Hong Q, Lebrilla CB, Leiserowitz GS, et al. Multiple Reaction Monitoring for the Quantitation of Serum Protein Glycosylation Profiles: Application to Ovarian Cancer. J Proteome Res. 2018 Jan 5;17(1):222–33.

18. Ruhaak LR, Kim K, Stroble C, Taylor SL, Hong Q, Miyamoto S, et al. Protein-Specific Differential Glycosylation of Immunoglobulins in Serum of Ovarian Cancer Patients. J Proteome Res. 2016;15(3):1002–10.

19. Saldova R, Wormald MR, Dwek RA, Rudd PM. Glycosylation changes on serum glycoproteins in ovarian cancer may contribute to disease pathogenesis. Dis Markers. 2008 Jan 1;25(4– 5):219–32.

20. Dhar, C., Ramachandran, P., Xu, G. et al. Diagnosing and staging epithelial ovarian cancer by serum glycoproteomic profiling. Br J Cancer 130, 1716–1724 (2024).

21. Wu Z, Serie D, Xu G, Zou J. PB-Net: Automatic peak integration by sequential deep learning for multiple reaction monitoring. J Proteomics. 2020 Jul;223:103820.

22. Luo, Robert; Boeras, Debi; Broyles, Laura N.; Fong, Youyi; …; Doherty, Meg MD; Vojnov, Lara, et al. Use of an Indeterminate Range in HIV Early Infant Diagnosis: A Systematic Review and Meta-Analysis. JAIDS Journal of Acquired Immune Deficiency Syndromes. 2019 Nov 1;82(3):p 281–286.

23. Alix-Panabières C, Marchetti D, Lang JE. Liquid biopsy: from concept to clinical application. Sci Rep. 2023 Dec 7;13(1):21685.

24. Pappas L, Adalsteinsson VA, Parikh AR. The emerging promise of liquid biopsies in solid tumors. Nat Cancer. 2022 Dec;3(12):1420–2.

25. Connal S, Cameron JM, Sala A, Brennan PM, Palmer DS, Palmer JD, et al. Liquid biopsies: the future of cancer early detection. J Transl Med. 2023 Feb 11;21(1):118.

26. Peracaula R, Sarrats A, Rudd PM. Liver proteins as sensor of human malignancies and inflammation. PROTEOMICS – Clin Appl. 2010;4(4):426–31.

27. An HJ, Miyamoto S, Lancaster KS, Kirmiz C, Li B, Lam KS, et al. Profiling of Glycans in Serum for the Discovery of Potential Biomarkers for Ovarian Cancer. J Proteome Res. 2006 Jul 1;5(7):1626–35.

28. Hua S, Williams CC, Dimapasoc LM, Ro GS, Ozcan S, Miyamoto S, et al. Isomer-specific chromatographic profiling yields highly sensitive and specific potential N-glycan biomarkers for epithelial ovarian cancer. J Chromatogr A. 2013 Mar 1;1279:58–67.

29. Kim K, Ruhaak LR, Nguyen UT, Taylor SL, Dimapasoc L, Williams C. Evaluation of Glycomic Profiling as a Diagnostic Biomarker for Epithelial Ovarian Cancer. Cancer Epidemiol Biomark Prev. 2014;1;23(4):611–21.

30. Dědová T, Braicu EI, Sehouli J, Blanchard V. Sialic Acid Linkage Analysis Refines the Diagnosis of Ovarian Cancer. Front Oncol [Internet]. 2019;9. Available from: https://www.frontiersin.org/journals/oncology/articles/10.3389/fonc.2019.00261

31. Qian, L., Zhu, J., Xue, Z. et al. Proteomic landscape of epithelial ovarian cancer. Nat Commun 15, 6462 (2024). 10.1038/s41467-024-50786-z

32. Jin, Y., Wang, W., Wang, Q. et al. Alpha-1-antichymotrypsin as a novel biomarker for diagnosis, prognosis, and therapy prediction in human diseases. Cancer Cell Int 22, 156 (2022). 10.1186/s12935-022-02572-4

33. Darwish NM, Al-Hail MK, Mohamed Y, Al Saady R, Mohsen S, Zar A, Al-Mansoori L, Pedersen S. The Role of Apolipoproteins in the Commonest Cancers: A Review. Cancers (Basel). 2023 Nov 24;15(23):5565. doi: 10.3390/cancers15235565.

34. Dieplinger H, Ankerst DP, Burges A, Lenhard M, Lingenhel A, Fineder L, Buchner H, Stieber P. Afamin and apolipoprotein A-IV: novel protein markers for ovarian cancer. Cancer Epidemiol Biomarkers Prev. 2009 Apr;18(4):1127–33. doi: 10.1158/1055-9965.EPI-08-0653. Epub 2009 Mar 31..

35. Ivanova TI, Klabukov ID, Krikunova LI, Poluektova MV, Sychenkova NI, Khorokhorina VA, Vorobyev NV, Gaas MY, Baranovskii DS, Goryainova OS, Sachko AM, Shegay PV, Kaprin AD, Tillib SV. Prognostic Value of Serum Transferrin Analysis in Patients with Ovarian Cancer and Cancer-Related Functional Iron Deficiency: A Retrospective Case-Control Study. J Clin Med. 2022 Dec 12;11(24):7377. doi: 10.3390/jcm11247377.

36. Lin Y, Bian L, Zhu G, Zhang B. Vitronectin promotes proliferation and metastasis of cervical cancer cells via the epithelial-mesenchymal transition. Front Oncol. 2024 Dec 9;14:1466264. doi: 10.3389/fonc.2024.1466264.

